# Scrutinizing the heterogeneous spreading of COVID-19 outbreak in Brazilian territory

**DOI:** 10.1101/2020.06.05.20123604

**Authors:** Rafael M. da Silva, Carlos F.O. Mendes, Cesar Manchein

## Abstract

After the spread of COVID-19 out of China, the evolution of the pandemic shows remarkable similarities and differences among countries across the world. Eventually, such characteristics are also observed between different regions of the same country. Herewith, we study the heterogeneous spreading of the confirmed infected cases and deaths by the COVID-19 until May 30th, 2020, in the Brazilian territory, which has been seen as the current epicenter of the pandemic in South America. Our first set of results is related to the similarities and it shows that: (i) a power-law growth of the cumulative number of infected people is observed for federative units of the five regions of Brazil; and (ii) the Distance Correlation (DC) calculated between the time series of the most affected federative units and the curve that describes the evolution of the pandemic in Brazil remains about 1 in most of the time, while such quantity calculated for the federative units with a low incidence of newly infected people remains about 0.95. In the second set of results, we focus on the heterogeneous distribution of the confirmed cases and deaths, which is demonstrated by the fact that only three regions concentrate 92% of the cases. By applying the epidemiological SIRD model we estimated the effective reproduction number *ℛ_e_* during the pandemic evolution and found that: (i) the mean value of *ℛ_e_* for the eight most affected federative units in Brazil is about 2; (ii) the current value of *ℛ_e_* for Brazil is greater than 1, which indicates that the epidemic peak is far; and (iii) Ceará was the only federative unit for which the current *ℛ_e_* < 1. Based on these findings, we projected the effects of increase or decrease the effective reproduction number and concluded that if the value of *ℛ_e_* increases 20%, not only the peak might grow at least 40% but also its occurrence might be anticipated, which hastens the collapse of the public health care system. In all cases, to keep the effective reproduction number 20% below the current one can save thousands of people in the long term.

---

June 5, 2020

## 1 Introduction

According to the World Health Organization (WHO), the Coronavirus Disease 2019 (COVID-19) is provoked by a pathogen agent known as Severe Acute Respiratory Syndrome Coronavirus 2 (SARS-CoV-2) [1]. Since the identification of SARS-CoV-2 in Wuhan, China, in December 2019, the pathogen kept spreading worldwide. One of the most remarkable characteristics of COVID-19 is its high infectivity, resulting in a global pandemic. Nowadays, tasks like protecting the people from the impacts of COVID-19 pandemic and the global economy are considered two major challenges. In this scenario, questions regarding the occurrence of the COVID-19 outbreak peak, how long it will persist, and how many people will be eventually infected are guiding a lot of studies.

Around the world, the battle against the COVID-19 spreading is based on several research approaches, which include treatments and preventive actions. However, to control the pandemic is found to be very challenging due to the following reasons: (i) it causes typical flu-like symptoms in human carriers; (ii) the human-to-human transmission via asymptomatic individuals; and (iii) the absence of proper clinical protocols (vaccine, drugs, and concrete ideas about the immunological response, for instance). In this complex scenario, an important research approach used to study the dynamics of the COVID-19 is the real time-series analysis, that has been extensively applied to obtain information about the evolution of the pandemic (infection, recovery, and death rates, for instance) [2, 3, 4, 5]. On the other hand, the uncertainty of the available official data, particularly related to the true number of infectious people, may lead to ambiguous results and inaccurate forecasts, as pointed out by other researchers [6] (see also references therein). For this reason, to explore the dynamics of COVID-19 by using combined approaches may be a necessary strategy to avoid misleading results.

Other important approaches used to study the evolution of COVID-19 are the epidemiological models like SIRD (Susceptible-Infected-Recovered-Dead) [3, 5, 7], SEIR (Susceptible-Exposed-Infected-Recovered) [8, 9, 10], and its modified versions [11, 12, 13, 14, 15, 16], which are fundamental tools to understand the evolution of the pandemic and to plan effective control strategies. One of the most important characteristics of these models is the quantity used to measure the transmission potential of the disease: the basic reproduction number *ℛ*_0_, which gives the number of secondary infected individuals generated by a primary infected in a population where all individuals are susceptible to infection. While for values *ℛ*_0_ < 1 the number of newly cases decreases exponentially, for 1 *< R*_0_ *< ∞* it increases exponentially [17, 18].

In recent studies about the COVID-19 outbreak, it was found a power-law growth of the cumulative number of infected individuals [11, 12, 13, 19, 20], which might be typical of small-world networks [21], and possibly is related to fractal kinetics and graph theory [22]. Such behavior seems to be general since the real time-series of countries from four distinct continents were fitted by power-law curves until March 27^th^[11]. Besides that, by calculating the Distance Correlation (DC) [23, 24, 25, 26] between the time series of these countries, it was possible to demonstrate that such data are highly correlated, which suggests a universal characteristic of the virus spreading.

Several studies have focused on the contagion scenarios in China [4, 5], Europe [5, 11, 12, 16], and United States of America (USA) [11, 12], due to the trajectory followed by the COVID-19 outbreak. As far as we know, there are only a few studies analyzing the dynamics of the COVID-19 in Brazil [27, 28, 29], which has the largest cumulative numbers of infected cases and deaths of South America. For this reason, we decided to investigate the dynamics of the COVID-19 in Brazil, which is the home of more than 210 million people, the world’s fifth-largest country by area, and currently the world’s eighth-largest economy. Another reason to study the dynamics of the COVID-19 outbreak in Brazil is due to the remarkable heterogeneity in the distribution of the infected people across the Brazilian territory. Brazil is composed of 26 federative units (or states) and one Federal District, which are distributed in five large regions: South, Southeast, Central-West, Northeast, and North. Besides the intrinsic differences between these regions, as the weather conditions (temperature and the duration of the raining seasons, for instance), cultural behaviors, and demographic density of people, the Human Development Indexes (HDI) are completely different if compared between the federative units. Interestingly, until May 30^th^, 32% of the cumulative number of infected cases and 44% of the deaths reported in Brazil due to COVID-19 happened in São Paulo (SP) and Rio de Janeiro (RJ), which represent the two largest economic units of the country and also concentrate a significant part of the population.

To scrutinize the heterogeneity of the COVID-19 spreading in Brazil, we propose to analyze the real time-series of all the federative units. Our main findings show that: (i) the cumulative number of infected people grows according to a power-law in all regions of Brazil; and (ii) the DC calculated between the time series of the most affected federative units and the time series of Brazil remains about 1 in most of the time, indicating a high similarity between the spread of the disease across such localities. Besides that, we used the SIRD model to estimate the effective reproduction number *ℛ_e_* and to project some scenarios based on its current value. A better and a worse scenario for the cumulative and active number of infected people, as well as for the number of dead people, were simulated and our findings corroborate the relevance of keeping the value of *ℛ_e_* the most next of 1 as possible. Additionally, the current value of the effective reproduction number reveals the epidemic stage and, in this way, may be used to indicate the right moment to act through containment measures.

The manuscript is organized as follows. Section 2 is designated to introduce an explanation about the real time-series collected and to define the methods applied here. Also, we explain the method to compute the DC between the time series and introduce the SIRD model formed by four Ordinary Differential Equations (ODEs). Section 3 discusses the numerical results obtained by fitting the real time-series, by calculating the DC between such data, and by applying the SIRD model to project different scenarios for the most affected Brazilian federative units. Finally, Section 4 summarizes our results and suggests some directions for future research studies.

## 2 Data and methods

This section is devoted to introducing the basic information about the real time-series and the methods used to explore the dynamics of COVID-19 spreading in the five Brazilian regions.

### 2.1 Real time-series

The real time-series refers to the cumulative number of confirmed cases of COVID-19 and deaths until May 30^th^, 2020, reported in the 27 federative units and also for the compiled results that form the time series of Brazil. Fundamental information about federative units, like their geographical position in Brazilian territory, the total population, the date of the first case, the cumulative number of confirmed cases, and deaths caused by the COVID-19 are detailed in Table 1. The time series were collected from the official website [30] that retrieves the daily information about COVID-19 cases from all 27 Brazilian State Health Offices, gather them, and make it publicly available. Among the 27 federative units, we focused our numerical investigations on 10 representative cases (two of each region) with a high incidence of COVID-19 cases: São Paulo (SP), Rio de Janeiro (RJ), Ceará (CE), Pernambuco (PE), Amazonas (AM), Pará (PA), Santa Catarina (SC), Paraná (PR), Federal District (FD) and Goiás (GO), which concentrate *∼*69% of the cumulative number of confirmed cases and *∼*82% of deaths. The time-series period of each state varies, once each state counts since the day of its first documented case until the day of the last report.

**Table 1.**
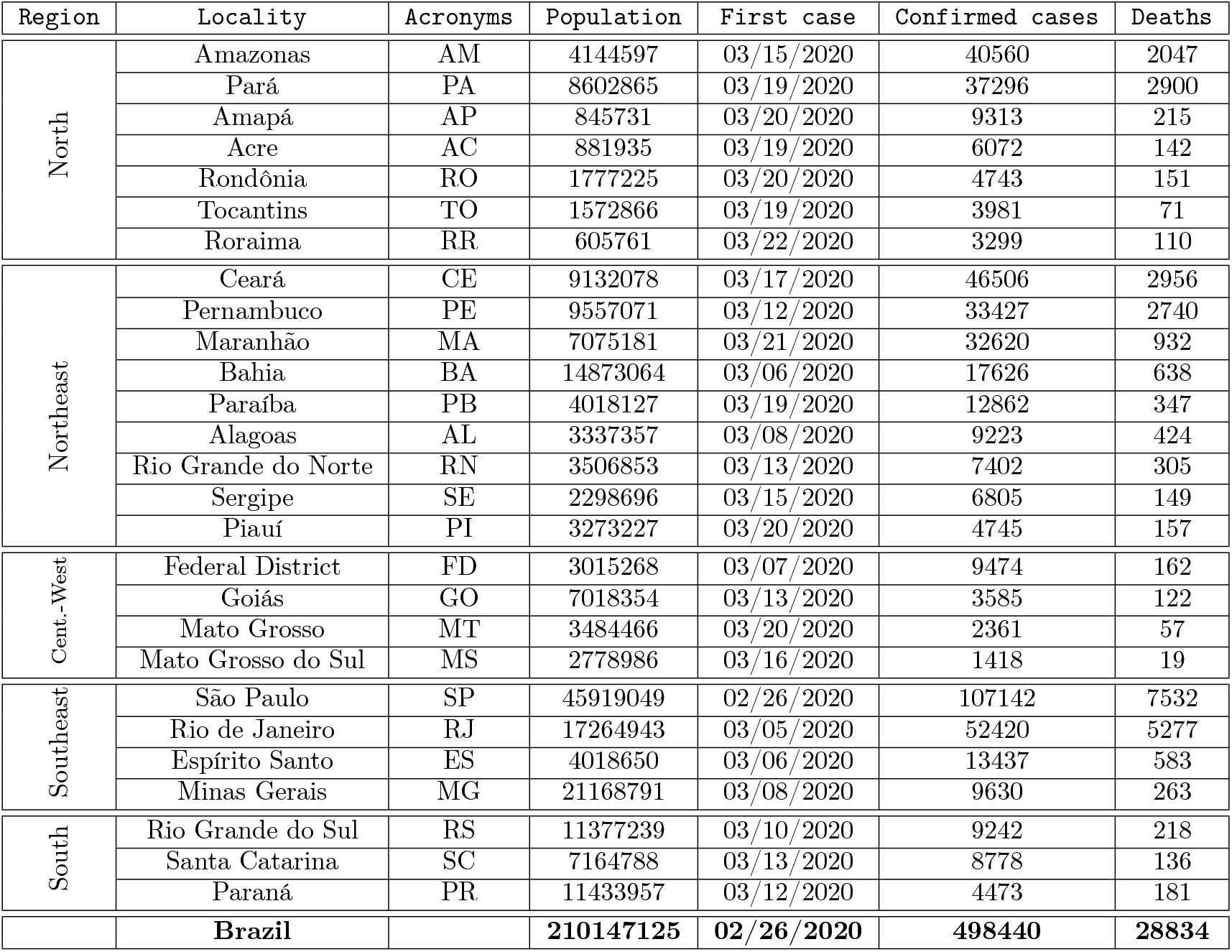
Data for the 27 federative units and for Brazil related to COVID-19 outbreak until May 30th [30]. Inside the block of each region, the federative units are listed in decreasing order of the number of confirmed infected cases.

| Region | Locality | Acronyms | Population | First case | Confirmed cases | Deaths |
| --- | --- | --- | --- | --- | --- | --- |
| North | Amazonas | AM | 4144597 | 03/15/2020 | 40560 | 2047 |
|  | Pará | PA | 8602865 | 03/19/2020 | 37296 | 2900 |
|  | Amapá | AP | 845731 | 03/20/2020 | 9313 | 215 |
|  | Acre | AC | 881935 | 03/19/2020 | 6072 | 142 |
|  | Rondônia | RO | 1777225 | 03/20/2020 | 4743 | 151 |
|  | Tocantins | TO | 1572866 | 03/19/2020 | 3981 | 71 |
|  | Roraima | RR | 605761 | 03/22/2020 | 3299 | 110 |
| Northeast | Ceará | CE | 9132078 | 03/17/2020 | 46506 | 2956 |
|  | Pernambuco | PE | 9557071 | 03/12/2020 | 33427 | 2740 |
|  | Maranhão | MA | 7075181 | 03/21/2020 | 32620 | 932 |
|  | Bahia | BA | 14873064 | 03/06/2020 | 17626 | 638 |
|  | Paraíba | PB | 4018127 | 03/19/2020 | 12862 | 347 |
|  | Alagoas | AL | 3337357 | 03/08/2020 | 9223 | 424 |
|  | Rio Grande do Norte | RN | 3506853 | 03/13/2020 | 7402 | 305 |
|  | Sergipe | SE | 2298696 | 03/15/2020 | 6805 | 149 |
|  | Piauí | PI | 3273227 | 03/20/2020 | 4745 | 157 |
| Cent.-West | Federal District | FD | 3015268 | 03/07/2020 | 9474 | 162 |
|  | Goiás | GO | 7018354 | 03/13/2020 | 3585 | 122 |
|  | Mato Grosso | MT | 3484466 | 03/20/2020 | 2361 | 57 |
|  | Mato Grosso do Sul | MS | 2778986 | 03/16/2020 | 1418 | 19 |
| Southeast | São Paulo | SP | 45919049 | 02/26/2020 | 107142 | 7532 |
|  | Rio de Janeiro | RJ | 17264943 | 03/05/2020 | 52420 | 5277 |
|  | Espírito Santo | ES | 4018650 | 03/06/2020 | 13437 | 583 |
|  | Minas Gerais | MG | 21168791 | 03/08/2020 | 9630 | 263 |
| South | Rio Grande do Sul | RS | 11377239 | 03/10/2020 | 9242 | 218 |
|  | Santa Catarina | SC | 7164788 | 03/13/2020 | 8778 | 136 |
|  | Paraná | PR | 11433957 | 03/12/2020 | 4473 | 181 |
|  | **Brazil** |  | **210147125** | **02/26/2020** | **498440** | **28834** |

### 2.2 Distance Correlation (DC)

The Distance Correlation (DC) is a statistical measure of the dependence between random vectors based on Euclidean distances [23] and is derived from quantities such as *variance* and *covariance of distance*. DC assumes null values if, and only if, the random vectors are completely independent, and can be defined from random vectors in arbitrary dimensions. For example, assuming that *l* and *m* are positive integers, the vectors can be defined as *X ∈* ℝ^*l*^and *Y ∈* ℝ^*m*^. In this case, the *distance covariance* between random vectors *X* and *Y* is defined by

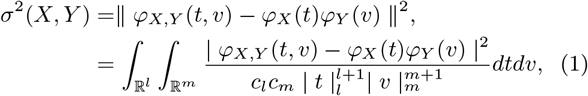

where *c_l_* and *c_m_* are constants, ║ · ║ is the Euclidean norm, *ϕ_X_*(*t*) and *ϕ_Y_* (*v*) are characteristic functions of *X* and *Y*, respectively, and *ϕ_X,Y_* (*t, v*) is the joint characteristic function. The joint characteristic function under the independence of two random vectors leads to the property *σ*^2^(*X, Y*) = 0 if, and only if, *X* and *Y* are independent, that is, *ϕ_X,Y_* (*t, v*) = *ϕ_X_* (*t*)*ϕ_Y_* (*v*) for all *t ∈* ℝ^*l*^and *v ∈* ℝ^*m*^. We can obtain similarly the *distance variance*, given by

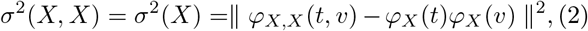

which is analogous for *σ*^2^(*Y*). Therefore, the DC coefficient between random vectors is defined by

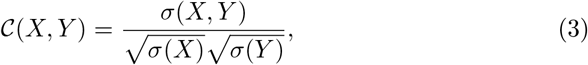

and such statistical measure is contained in the interval [0, 1].

#### 2.2.1 Computational procedure to compute the DC

The computational method defined here will be used to analyze statistically the real data of the Brazilian federative units regarding the COVID-19 outbreak. Consider a joint sample (*X, Y*) = *{*(*X*_1_*, Y*_1_), (*X*_2_*, Y*_2_),…, (*X_N_, Y_N_*)*}* defined by *i, j* = 1,…, *N*, with *X ∈* ℝ^*l*^, *Y ∈* ℝ^*m*^and *N ≥* 2. Thus, the *empirical DC coefficient* for a joint sample (*X, Y*) is given by

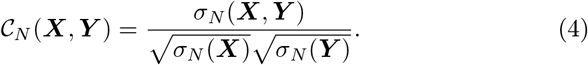

The *empirical distance covariance* for (*X, Y*) is given by

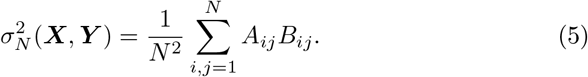

To calculate numerically the distance covariance by using Eq. (5) is much simpler than by using Eq. (1), although both equations describe the same quantity. For details about the equivalence between such definitions, we suggest the Ref. [23].

For a sample *X*, the *empirical distance variance* is given by

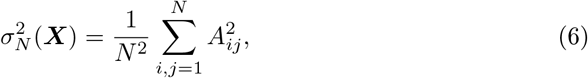

and for a sample *Y*,

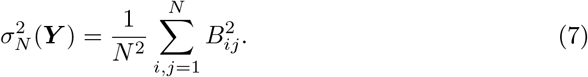

The matrix *A_ij_* is obtained from *X*:

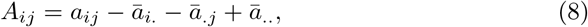

where 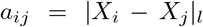 is the Euclidean norm of the distance between the pairs of elements of the sample, 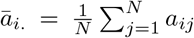 is the arithmetic mean of the rows, 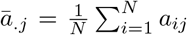 is the arithmetic mean of the columns, and 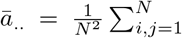 is the general mean. Similarly, the matrix *B_ij_* is obtained from *Y*.

It is important to mention that, to obtain the DC by applying the procedure described above, we need only the samples *X* and *Y*. This can be quite relevant when it comes to experimental data. In Eq. (4) it is possible to verify that *𝒞_N_*(*X, Y*) is independent of the scale since the samples *X* and *Y* can be multiplied by a real number and *𝒞_N_*(*X, Y*) remains unchanged. This is an important feature of the DC coefficient which allows us to obtain relevant information even if data with different magnitudes are compared. Furthermore, it is important to mention that, if all elements of a sample *X* or *Y* are identical, then we have *σ_N_*(*X*)*σ_N_*(*Y*) = 0 and therefore *𝒞_N_*(*X, Y*) = 0 as shown in [23].

### 2.3 The SIRD model

Since the pioneering work of Kermack and McKendrick [31], mathematical models have been developed to describe infectious disease dynamics and to help us to analyze the efficiency of preventive measures that could be adopted to control infections spread. Containment strategies such as social distance, which includes mask-wearing, quarantine, and other actions, are essential to avoid the uncontrolled increase in the number of infected people and the collapse of the health care systems.

In this work, we used the well-known SIRD model to reproduce the realistic data, to estimate some important epidemiological parameters, and to study the effects of containment measures around the different regions of Brazil. In this model, the total population *N* is divided into subgroups of Susceptibles (*S*), Infected (*I*), Recovered (*R*), and Dead (*D*) people. For all time *t, N* = *S* + *I* + *R* + *D*. The ODEs that describe the dynamics of such groups are the following [31, 32]:

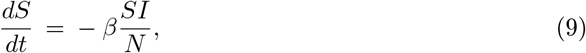

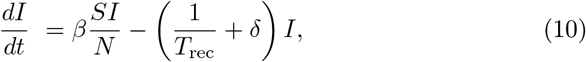

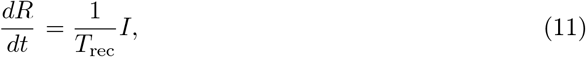

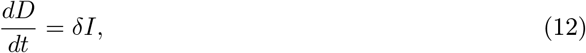

where *β, T*_rec_, and *δ* are “effective” parameters, once they are based on the reported confirmed cases and deaths. Fig. 1 shows a schematic representation of the SIRD model with the relationship between variables and parameters. The parameter *β* is the infection rate, which reflects the probability per unit time that a susceptible individual becomes infectious when entering in contact with an infected person. The term 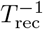 is the recovery rate, with *T*_rec_ being the infection duration or the recovery time. Finally, *δ* is the death rate, which means the portion of people that die per unit time in comparison with the number of active infected people. The initial conditions are [*S*(*t*_0_)*, I*(*t*_0_)*, R*(*t*_0_)*, D*(*t*_0_)], and the population at each time is given by *N*(*t*) = *N*(*t*_0_) *− D*(*t*), being *N*(*t*_0_) the initial population described in Table 1. To update the population is important for future projections, once the number of deaths *D*(*t*) could be significant when compared to *N*(*t*). In our simulations, we set *R*(*t*_0_) = 0 and *I*(*t*_0_)*, D*(*t*_0_) were obtained from the real time-series. We considered that the initial number of susceptible people is given by *S*(*t*_0_) = *N*(*t*_0_) *− I*(*t*_0_) − *R*(*t*_0_) *− D*(*t*_0_). In addition to these equations, we compute the cumulative number of confirmed cases *C*(*t*) of COVID-19 by the relation *C*(*t*) = *I*(*t*)+*R*(*t*)+ *D*(*t*).

**Figure 1.**
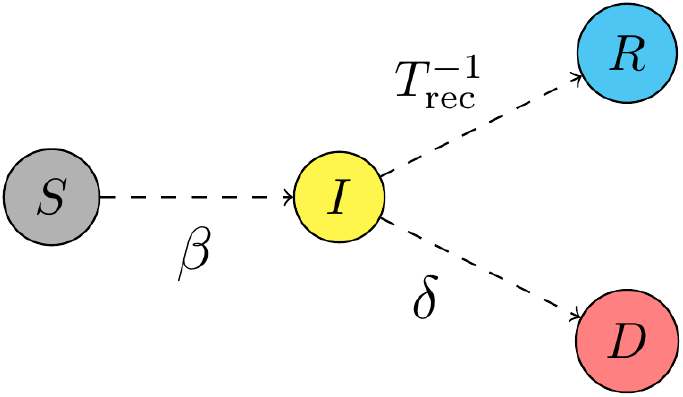
Schematic representation of the SIRD model highlighting its variables and the role of the parameters connecting the variables.

The values of *β, T*_rec_, and *δ* depend on several circumstances and must differ significantly in different regions and different periods of the pandemic. Besides, to obtain reliable values for these parameters in each locality, we propose to adjust the parametric set (*β, T*_rec_*, δ*) using the evolution of the real data. To start the fitting, first of all, we divided the time series in epidemiological weeks (EPI weeks), which are, by international convention, count from Sunday to the next Saturday. With this split, we intend to find the best combination of *β*(*k*), *T*_rec_(*k*), and *δ*(*k*) that describes the evolution of the pandemic in each EPI week number *k*. Moreover, the number of data inside each EPI week satisfies the minimal number of 2*r* + 1 experiments with real data needed to adjust trusty *r* unknown parameters [33] (in our case, *r* = 3).

To define the parametric set in each EPI week, we performed simulations for all possible combinations obtained by varying *β ∈* [0.0, 0.5] with a step of 10^−2^, *T*_rec_ *∈* [10, 21] with a step of 10^−1^, and *δ ∈* [10^−4^, 3 *×* 10^−2^] with a step of 10^−4^. For each set (*β, T*_rec_*, δ*), we(i) calculated the Root Mean Square Error (RMSE) ∆_*C*_ between the cumulative number of infected people *C*(*t*) obtained by the model in each day and the real data; and (ii) calculated the RMSE ∆_*D*_ between the number of deaths *D*(*t*) obtained by the model in each day and the real data. The best parametric set is that for which the quantity 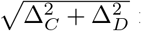 is minimized. We picked up the range 10–21 days for the recovery time because it covers a reliable estimate around the world [3, 7, 34]. On the other hand, in Brazil, a variation from 0.01% to 3% for the daily death rate *δ* is a satisfactory range [30, 35]. To obtain better results, the first day considered on the fitting is the next Sunday after the first death in each time series analyzed. For example, the first death in Brazil was confirmed on March 17^th^, the Tuesday belonging to the EPI week number 12. Then, we started adjusting the parametric set from March 22^nd^, the beginning of the EPI week number 13.

## 3. Results

### 3.1 Power-law grows

Figure 2 displays the data of the cumulative number of infected people by COVID-19 of ten representative federative units and of Brazil as a function of the days, using the linear scale. The analyzed federative units were chosen to represent all the five regions that compose the Brazilian territory (see Table 1 and the caption of Fig. 2). Each block that composes the Table 1 represents one Brazilian region, where the federative units are listed in decreasing order of the number of infected people. In Fig. 2, the black and brown continuous curves are the corresponding fitting curves *α* + *γt^µ^*, where *t* is the time given in days after the reported hundredth case, *α, γ* and *µ* are parameters. We choose to discard the initial data, regarding the days with less than 100 infected individuals, since in this way it was possible to capture the real tendency of the fitted curves and avoid misunderstandings due to poor statistics. The straight lines in each inset, plotted in logarithm scale, indicate power-law growth as previously observed for different countries in four continents [11, 12].

**Figure 2.**
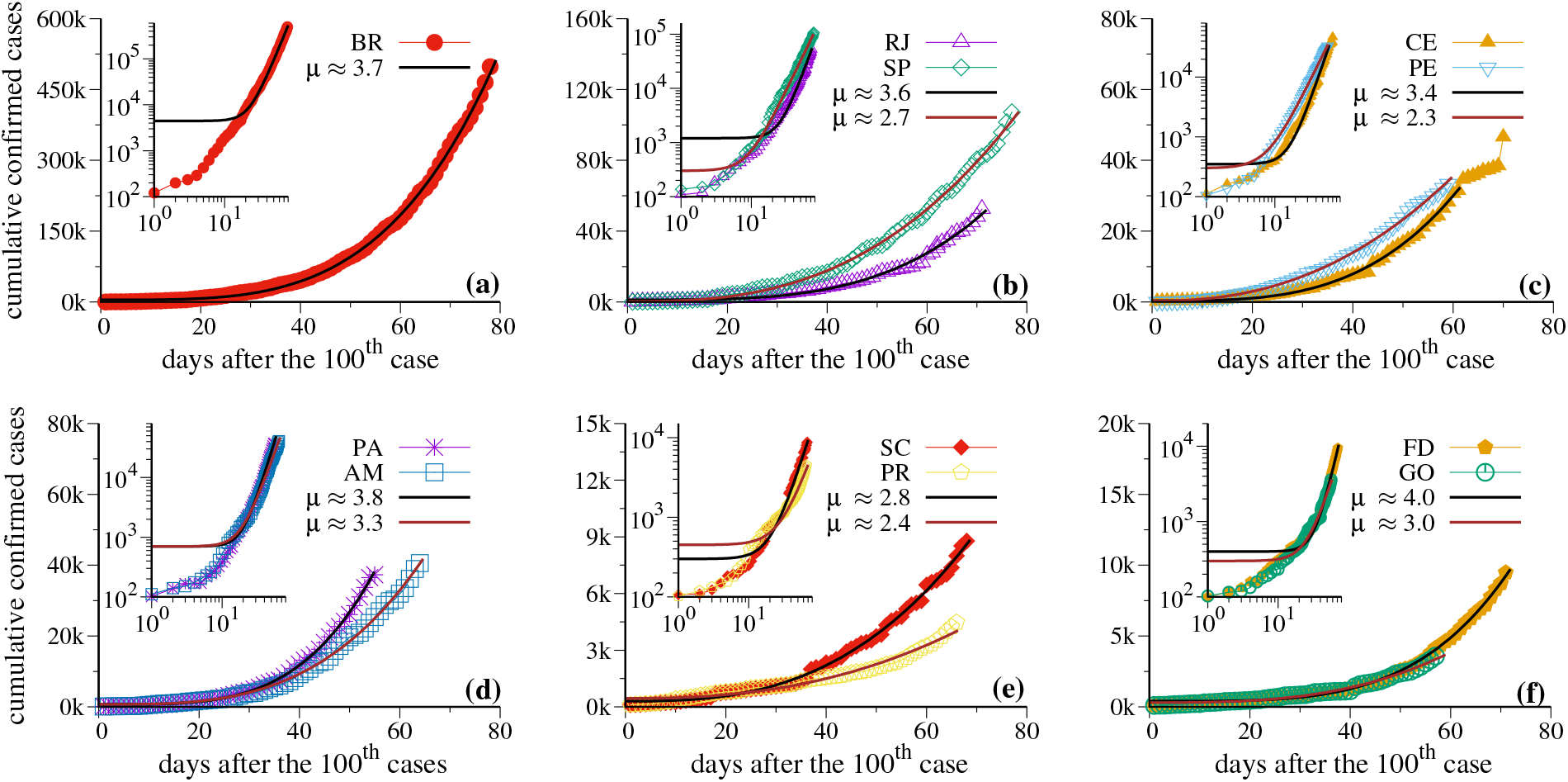
The cumulative number of confirmed cases of COVID-19 as a function of time for (a) Brazil, (b) SP and RJ, (c) CE and PE, (d) AM and PA, (e) SC and PR, (f) FD and GO. The same quantities are also plotted in logarithm scale in the insets. The black and brown continuous curves represent the function *α* + *γt^μ^* that fit the time-series, and the parameters *α, γ*, and *μ* for each locality are described in Table 2.

It is fundamental to understand the stage of the pandemic to take the right decision. This knowledge may be obtained through the contamination’s curves since they provide essential information about what is expected in the future and about what can be done to flatten the curves. Such information is encoded in the exponent *µ* that changes for distinct federative units. The complete fitting parameters are given in Table 2. Results in Table 2 are presented in the same order as plotted in Fig. 2. Through this figure, it is clear that the cumulative number of confirmed cases is still growing according to power-laws with different exponents *µ* in the most federative units as in the country. These differences may be caused by several factors as, for example, due to the size of the federative units, the heterogeneous concentrations of poor people, or the complex social and political organizations of them, that have applied different strategies in different stages of the contagious process. Nowadays, the administrative organization of Brazil gives to the federative units and municipalities some independence to adopt mitigation approaches, but these are limited by economic dependence on superior spheres. So, the tendency is to have similar approaches within federative units, although someones have obtained much better results than others. Interestingly, while PA and FD have exponents *µ* larger than Brazil, the other federative units have smaller exponents.

**Table 2.** Details about the parameters of the fitting curves for the power-law behavior *α* + *γt^μ^* shown in Fig. 2.

| Locality | $\alpha$ | $\gamma$ | $\mu$ |
| --- | --- | --- | --- |
| Brazil | 4450 | 0.040 | $3.743 \pm 0.022$ |
| RJ | 1200 | 0.011 | $3.600 \pm 0.064$ |
| SP | 300 | 0.052 | $2.699 \pm 0.022$ |
| CE | 350 | 0.008 | $3.367 \pm 0.037$ |
| PE | 300 | 2.428 | $2.340 \pm 0.023$ |
| PA | 700 | 0.008 | $3.849 \pm 0.032$ |
| AM | 700 | 0.052 | $3.259 \pm 0.033$ |
| SC | 300 | 0.066 | $2.783 \pm 0.035$ |
| PR | 450 | 0.129 | $2.441 \pm 0.080$ |
| FD | 400 | $2.892 \times 10^{-4}$ | $4.044 \pm 0.035$ |
| GO | 300 | 0.019 | $2.971 \pm 0.068$ |

Additionally, SP and Brazil present quite similar *μ* values, where *μ ∼* 3.6. This is not a coincidence, as discussed in detail in the next subsection.

The most desired behavior is that the exponent *μ* becomes smaller, leading to the flattening of the curves. Although, to achieve this scenario is not that easy. Besides CE, which seems to be stabilizing the epidemic spread, in all other federative units the growth remains strictly on the power-law fitting curve and *μ* essentially does not change in time. In Subsection 3.4.2, we discuss some possibilities to flatten the power-laws.

### 3.2 The DC between Brazil and its federative units

Figure 3 presents the DC coefficient *𝒞_N_*(*X, Y*) (plotted in colors) computed between the real time-series of Brazil (sample *{X}*) and each one of its 27 federative units (sample {*Y*}), considering the data for the cumulative number of cases in Fig. 3(a) and the number of deaths in Fig. 3(b). For simplicity, we now will use only the notation *C* for the DC coefficient. As mentioned in Subsection 2.2, the DC is a measure that analyzes how similar the shapes of the curves are to each other independently of the magnitude of such data. With this statistical analysis, we intend to find the federative units for which the time series is more correlated with the data of Brazil. Since *C* measures the “similarity” of such curves, with this information it is possible to know the states that are still following a power-law growth together with the country or are flattening the curve. The different colors in Fig. 3 correspond to distinct values for *𝒞* between the real time-series. According to the palette, the black region corresponds to the absence of data (ND), *i.e*., without reported infected people and deaths for the federative units, except for SP. The white and the gray regions mean a low value for DC, while the red, yellow, cyan and navy blue colors are related to high correlations.

In Fig. 3(a), all the federative units are shown from the left to the right in alphabetical order. On the vertical axis, the date is displayed since February 26^th^, the day when the first case of COVID-19 was reported in SP. In this figure, we can see that the time series of SP is highly correlated with the time series of Brazil since the first reported case, which is indicated by the long vertical navy blue stripe (*𝒞 ∼* 1). This result was expected since the power-law growth of the cumulative number of infected people for both Brazil and SP shows a very similar exponent, with *μ ∼* 3.6 (see Table 2 and Fig. 2). Similarly, we observe that the time series of BA, CE, and RJ were also highly correlated in most of the time with the data of Brazil, which means that such federative units also showed a power-law increasing of the cumulative number of infected people, how can be observed in Fig. 2.

On the other hand, it is interesting to note that for PR, in the period from April 22^nd^ to May 20^th^, the coefficient *𝒞* suffers a slight decay, remaining around 0.95. This value reflects the oscillations in the time series of PR, which can be seen in Fig. 2(e). SE and TO, two federative units with a relatively low incidence of infected people, have presented an evident similarity in the behavior of the DC coefficient in time. For both states, the value of *𝒞* presented oscillations around 0.90 and 0.98 during the period considered.

Another important achievement of the DC coefficient is to identify a possible change in the trend of one time series. Fig. 2(c) shows that CE is apparently on the way of flattening the curve, which no longer follows a power-law growth. Since this new behavior is recent, the change in the value of *𝒞* is slight and can not be depicted by the color scale in Fig. 3. For this reason, we plotted the time series of Brazil and CE in Fig. 4(a) and the value of *𝒞* calculated between such data in Fig. 4(b). In this figure, it is possible to see that, while both time series follow a power-law growth, the DC coefficient increases until 1. However, once that CE starts flattening its curve and Brazil continues growing up, *𝒞* starts to decrease.

**Figure 3.**
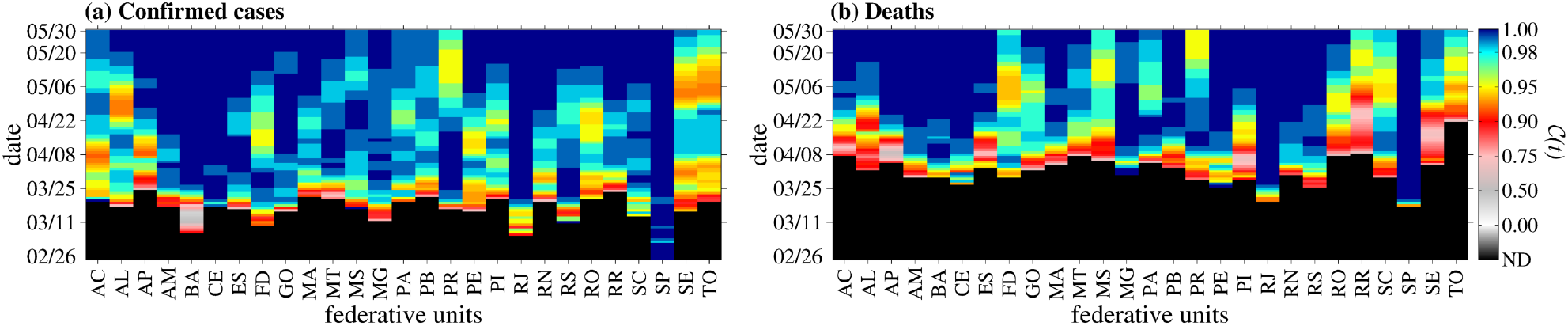
DC coefficient (*𝒞*) plotted in colors for the cumulative number of (a) confirmed cases and (b) deaths. Analyzing these panels, it is possible to see that BA, CE, RJ, and SP showed a behavior similar to Brazil in most of the time, which is indicated by the long vertical navy blue stripes. On the other hand, the value of *𝒞* for states that show a different behavior is lower (about 0.95), as is possible to see for PR, SE, and TO, for instance.

**Figure 4.**
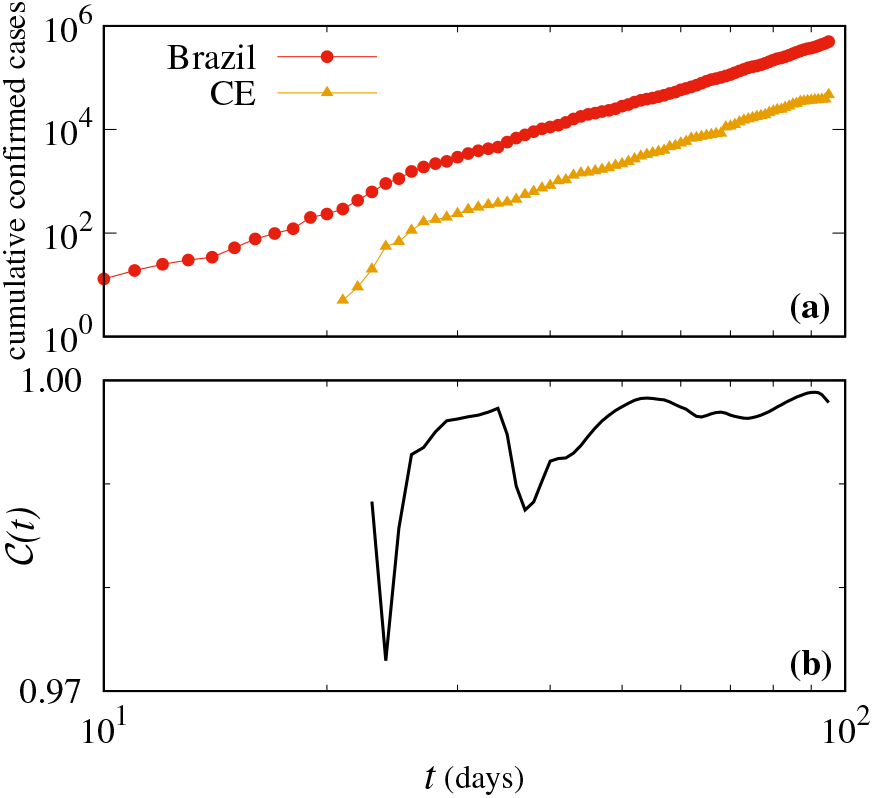
(a) The time series of the cumulative number of confirmed cases of Brazil and CE and (b) the DC coefficient (*𝒞*) calculated between these data. On the horizontal axis, the days are count since the first case in Brazil.

In Fig. 3(b) we plotted the value of *𝒞* for the real time-series of the number of deaths caused by COVID-19. In this case, it is remarkable that the times series of SP and RJ, the two federative units with the greatest numbers of deaths, are highly correlated with the data of Brazil in most of the time. Besides that, it is also possible to note that AM, BA, and CE reached high values of *𝒞* from April 22^nd^.

In general, the oscillations presented by *𝒞*, which are consequences of the different behaviors of the data along the time, testify the heterogeneity of the COVID-19 spreading in the Brazilian territory. Such differences might be related to some reasons as, for example, the different containment measures adopted in each region, the lack of testing, underreported data, and others.

### 3.3. An estimate of the effective reproduction number

As explained in Section 1, the basic reproduction number *ℛ*_0_ is the average number of secondary infections originated from an infectious person considering a population where everyone is susceptible. However, this condition is strictly valid only when the pathogen is in the early days. With the evolution of time, the susceptible fraction of the population decreases, and the fundamental epidemiological quantity that determines the dynamical evolution of the pandemic is the *replacement number* or the *effective reproduction number ℛ_e_*(*t*), which is defined as the average number of newly infected individuals produced by a typical infective during the period of infectiousness [18]. This quantity captures changes in the pandemic evolution and the key aim of the government containment measures is to reduce *ℛ_e_*(*t*). If *ℛ_e_*(*t*) < 1, the incidence of new infections decreases, and the epidemic is controlled. On the other hand, if *ℛ_e_*(*t*) > 1, the number of newly infected people will continue to grow until the peak and, eventually, can decline due to the acquisition of immunity.

The effective reproduction number *ℛ_e_*(*t*) is a dynamical quantity, and by using the parameters obtained by the procedure described in Subsection 2.3 we can estimate the value of *ℛ_e_*(*k*) for each EPI week*k* using the relation

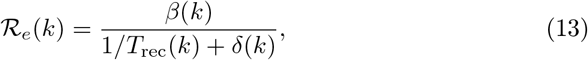

where *β*(*k*), *T*_rec_(*k*), and *δ*(*k*) is the best parametric combination for each EPI week. The effective reproduction number calculated for the data of Brazil is shown in Fig. 5.

**Figure 5.**
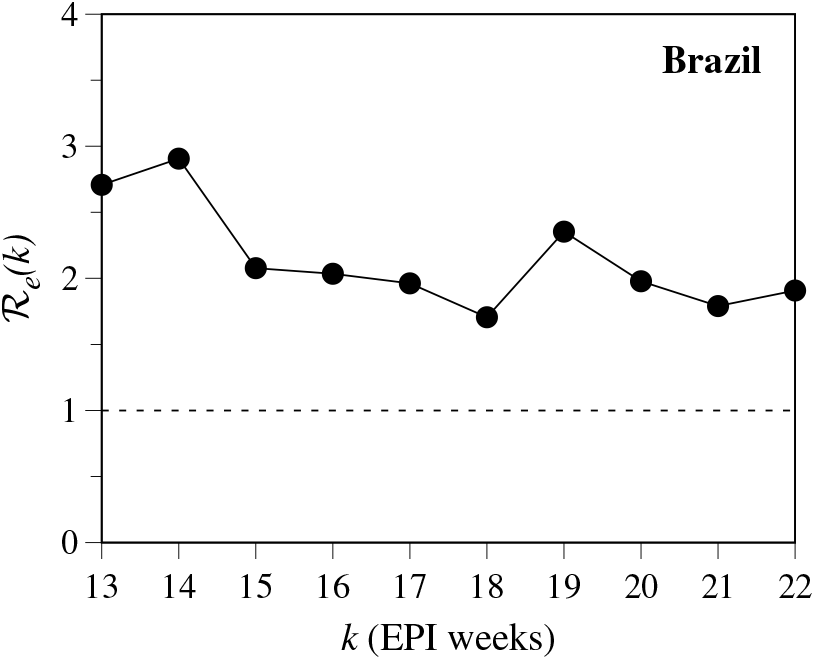
The effective reproduction number *ℛ_e_* estimated for each EPI week *k* for Brazil.

The first case of COVID-19 in Brazil was confirmed during the EPI week 9, and the value of the basic reproduction number *ℛ*_0_ is estimated in the range of 3–4, in line with estimates of transmissibility derived from Europe [35]. We can observe in Fig. 5 that *ℛ_e_*(13) = 2.7 and *ℛ_e_*(14) = 2.9, results obtained after some federative units had adopted the first containment measures during the EPI week 12. After these weeks, the effective reproduction number continued decreasing, reaching the value 1.7 in the EPI week 18. In early May, a relaxing on the social distance measures was observed in Brazil and the number of newly infected cases exploded. Our simulations demonstrate this increase in the value of *ℛ_e_*(*k*) in the EPI week number 19, when *ℛ_e_ >* 2 again. After this increase, the value of *ℛ_e_* remained below 2 in the last EPI weeks.

It is important to mention that there are other more precise methods to estimate the value of *ℛ_e_* (see, for example, the Refs. [35, 36, 37]). However, even though the SIRD model is rather crude, our simulations provide reliable values for the reproduction numbers and, more importantly, capture trends for such quantity. Besides the mean value of *ℛ_e_*(*k*), we display in Table 3 the mean values for the infection rate *β*(*k*), recovery time *T*_rec_(*k*), and death rate *δ*(*k*) for Brazil and for the eight federative units with the higher incidence of infected people, which are, in decreasing order: SP, RJ, CE, AM, PA, PE, MA, and BA. These mean values were calculated over all EPI weeks considered in our analysis. The value of *ℛ_e_*(22) obtained in the EPI week number 22 is also displayed since such quantity will be used in the next subsection to analyze the trends for *𝒞*(*t*) and *D*(*t*) and to simulate effects of increase or decrease the effective reproduction number on the pandemic evolution in Brazil and its federative units.

**Table 3.**
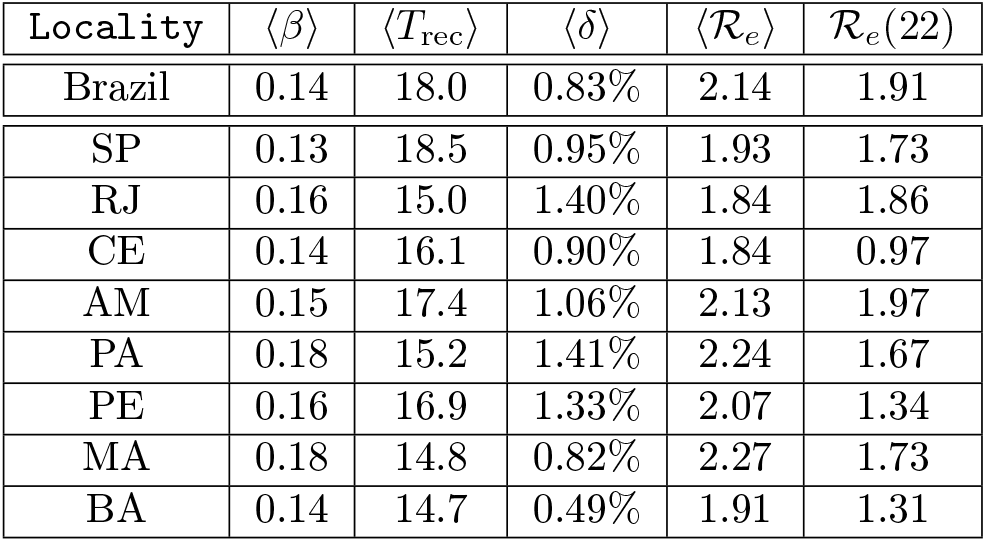
Table of mean values of the epidemiological parameters obtained by fitting the real time-series. The values of 〈*β*〉 and 〈*δ*〉 are given in days^−1^, 〈*T*_rec_〉 is given in days, and

| Locality | $\langle\beta\rangle$ | $\langle T_{\text{rec}}\rangle$ | $\langle\delta\rangle$ | $\langle\mathcal{R}_e\rangle$ | $\mathcal{R}_e(22)$ |
| --- | --- | --- | --- | --- | --- |
| Brazil | 0.14 | 18.0 | 0.83% | 2.14 | 1.91 |
| SP | 0.13 | 18.5 | 0.95% | 1.93 | 1.73 |
| RJ | 0.16 | 15.0 | 1.40% | 1.84 | 1.86 |
| CE | 0.14 | 16.1 | 0.90% | 1.84 | 0.97 |
| AM | 0.15 | 17.4 | 1.06% | 2.13 | 1.97 |
| PA | 0.18 | 15.2 | 1.41% | 2.24 | 1.67 |
| PE | 0.16 | 16.9 | 1.33% | 2.07 | 1.34 |
| MA | 0.18 | 14.8 | 0.82% | 2.27 | 1.73 |
| BA | 0.14 | 14.7 | 0.49% | 1.91 | 1.31 |

### 3.4. Simulating the effects of increase or decrease the effective reproduction number

In this section, we describe the results obtained by using the SIRD model to fit the real data and to project some scenarios. First, we analyzed the trends for the next days assuming that the effective reproduction number *ℛ_e_*(22) of the EPI week number 22 remains unchanged. Thereon, we study the effects of changing the effective reproduction number on quantities as *𝒞*(*t*), *D*(*t*), and *I*(*t*). Such variables are certainly affected by the recovery rate *T*_rec_, and also by the death rate *δ*. However, since these parameters are mostly changed by pharmacological interventions, in our simulations they were kept fixed and we increase or decrease the value of *β*(22), which is the quantity that reflects non-pharmacological containment measures as, for example, the social distance actions. With these simulations, we do not intend to preview an exact number of infected or dead people. Actually, we know that the epidemiological parameters are dynamical and change recurrently, which turns long-term projections a tough task. The results presented in the next subsections aim to demonstrate as a simple mathematical model can be used to describe the real data and to convince about the relevance of taking preventive measures as soon as possible to contain the virus spreading and to save lives.

#### 3.4.1 The cumulative number of cases and deaths

Figure 6 shows the real times-series for the cumulative number of confirmed cases of COVID-19 (blue circles) and deaths (black squares) for Brazil and for the eight federative units most affected by the pandemic. In this Figure, we also display the curves of *𝒞*(*t*) and *D*(*t*) obtained by integrating the equations of the model, represented by the dark blue and black continuous lines, respectively. We observe that, by adjusting the parameters *β, T*_rec_, and *δ* in each EPI week (represented by the gray rectangles in the background), it is possible to describe precisely the behavior of the real data for all localities analyzed. After to find the best parameter combination that fits the real data for each EPI week, the dark blue and black continuous curves describe the trend for *𝒞*(*t*) and *D*(*t*), respectively, assuming that the values *β*(22), *T*_rec_(22), and *δ*(22) of the EPI week number 22 will remain unchanged until the day 180 after the first case. Therefore, by Eq. (13), this assumption means to keep fixed the same effective reproduction number obtained in the EPI week 22, value that is shown in Table 3 for the localities studied.

**Figure 6.**
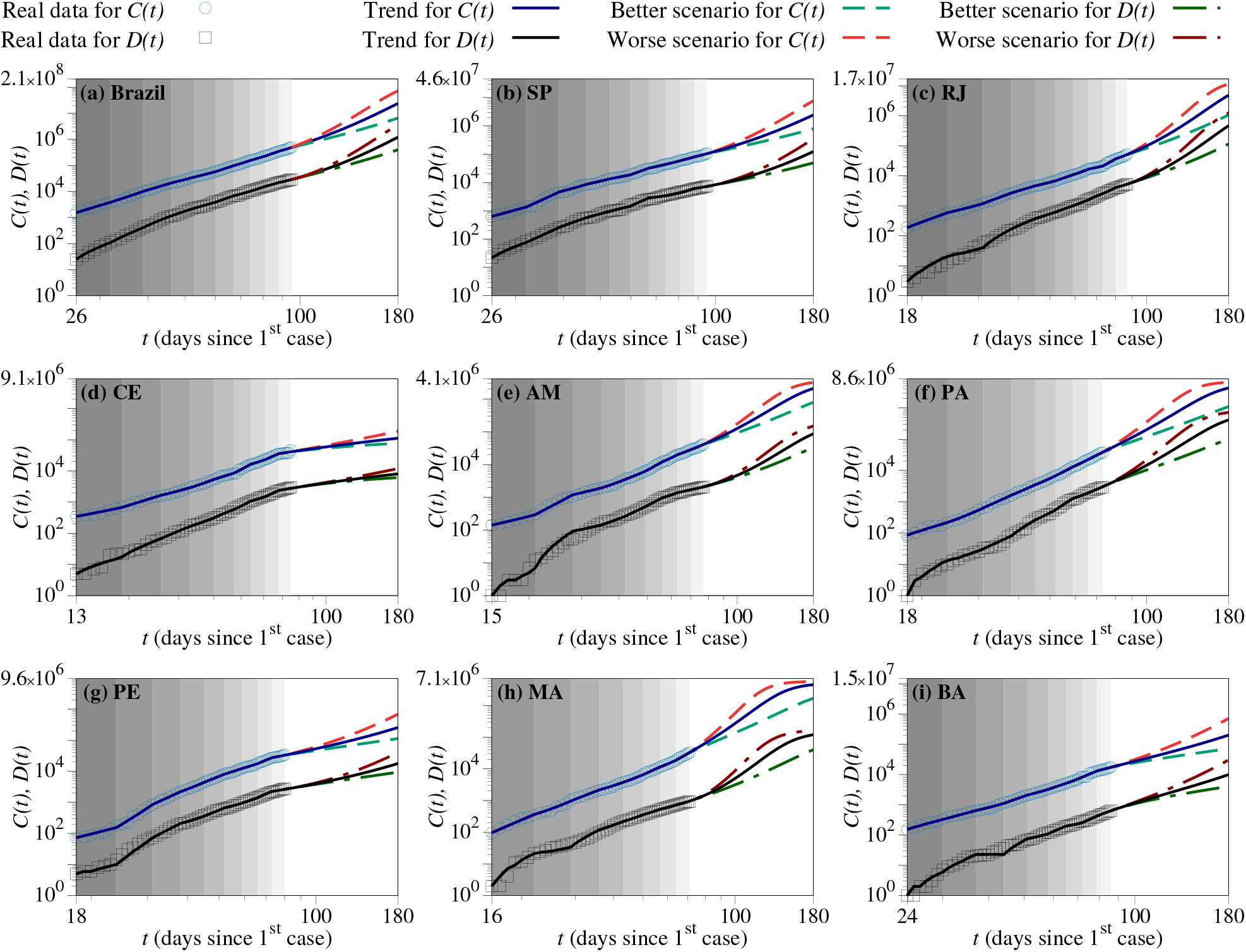
The cumulative number *C*(*t*) of confirmed cases of COVID-19 (blue circles) and deaths *D*(*t*) (black squares) for (a) Brazil, (b) SP, (c) RJ, (d) CE, (e) AM, (f) PA, (g) PE, (h) MA, and (i) BA. The dark blue and black continuous curves show the trends for *C*(*t*) and *D*(*t*), respectively, assuming that the effective reproduction number of the EPI week number 22 is kept fixed. A better scenario means to reduce in 20% the *ℛ_e_*(22), and this case is represented for *C*(*t*) by the light green dashed lines and for *D*(*t*) by the dark green dash-dotted lines. A worse scenario means to increase in 20% the *ℛ_e_*(22), and this case is represented for *C*(*t*) by the light red dashed lines and for *D*(*t*) by the dark red dash-dotted lines.

In Figs. 6(a), 6(b), and 6(c), we see the trend for Brazil, SP, and RJ, in this order. SP and RJ are the most populous federative units of Brazil and together they cover 32% of the total number of confirmed cases of the country. In these three cases, the effective reproduction number *ℛ_e_*(22) is greater than 1.7, and it is possible to note that the trend for *𝒞*(*t*) and *D*(*t*) is to keep increasing for several weeks. Besides these cases, we also highlight the examples of AM [Fig. 6(e)], PA [Fig. 6(f)], and MA [Fig. 6(h)], for which *ℛ_e_*(22) > 1.6. Our simulations show that, if the effective reproduction number does not change, about half of the initial population *N*(*t*_0_) of such federative units can be infected until the day 180 after the first case. It is possible for these three states since their population is considerably smaller than SP and RJ, for example (see Table 1). Besides that, according to

Table 3, these three federative units show the worst values of 〈*ℛ_e_*〉 over the EPI weeks considered.

Differently from the cases discussed above, PE [Fig. 6(g)] and BA [Fig. 6(i)] show a more attenuated increase for *C*(*t*) and *D*(*t*), which reflects the fact that their effective reproduction number in the EPI week 22 is about 1.3. The best result was obtained for CE, the case displayed in Fig. 6(d), for which *ℛ_e_*(22) < 1. In this case, the growth of the curves *C*(*t*) and *D*(*t*) is very slow, which means that the pandemic is under control if this scenario is kept.

Since the value of *ℛ_e_* depends on several events and can change daily, then we project some scenarios considering the increasing and the decreasing of such quantity. For this, we kept fixed *T*_rec_(22) and *δ*(22), and changed the value of *β*(22), summing and reducing 20% of its value, which means to increase or decrease in 20% the effective reproduction number. A better scenario was simulated by decreasing *ℛ_e_*(22) in 20%, and this case for *C*(*t*) is represented by the light green dashed curves, and for *D*(*t*) by the dark green dash-dotted curves. In all cases, to keep the effective reproduction number 20% below the current one can save thousands of people in the long term. On the other hand, a worse scenario takes place when increasing in 20% the current value of *ℛ_e_*, which is represented for *C*(*t*) by the light red dashed curves, and for *D*(*t*) by the dark red dash-dotted curves. This scenario increases both the number of confirmed cases and deaths for all localities.

#### 3.4.2. Flattening the curve of active infected individuals

Now, we will describe the results obtained by analyzing the number of active infected people *I*(*t*). This analysis is important since a huge number of simultaneously infected people certainly lead the health care systems to collapse. Figure 7 shows three curves for each locality: (i) the dark blue continuous curve describes the trend, obtained by keeping the value *ℛ_e_*(22) for the next days; (ii) the dark green dashed line describes a better scenario, obtained by decreasing *ℛ_e_*(22) in 20%; and (iii) the dark red dash-dotted line describes a worse scenario, obtained by increasing *ℛ_e_*(22) in 20%. Analyzing Fig. 7, we observe that increasing the value of the effective reproduction number in 20%, the peak of *I*(*t*) not only growths but also occurs earlier. On the other hand, decreasing *ℛ_e_*(22), the desired flattening of the curves of infected people is reached, which means to shrink the peak and to delay its occurrence.

**Figure 7.**
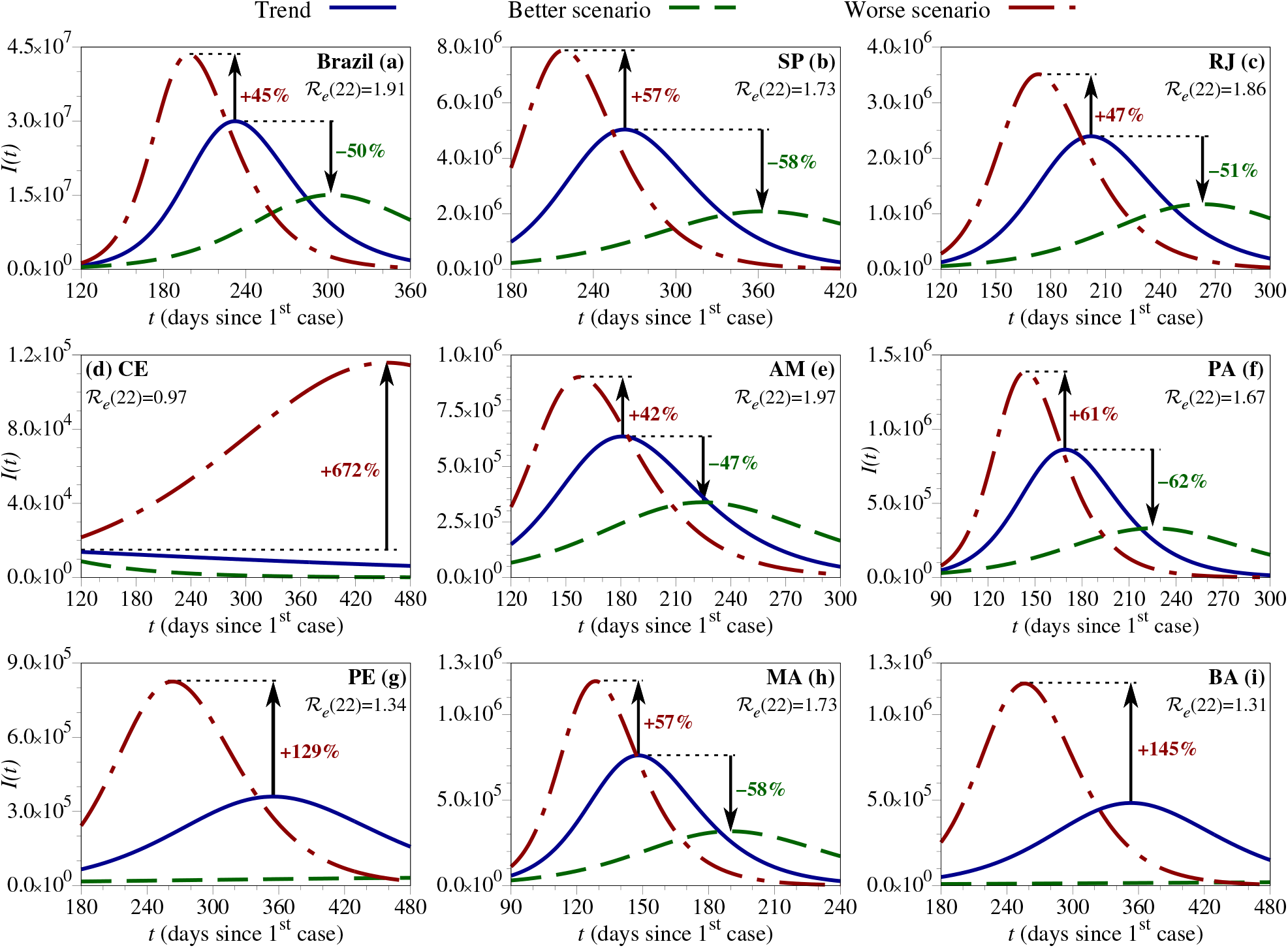
Projections for the number of active infected individuals *I*(*t*) obtained by keeping the value of *ℛ_e_*(22) (dark blue continuous curves), by decreasing in 20% the value of *ℛ_e_*(22) (dark green dashed curves), and by increasing in 20% the value of *ℛ_e_*(22) (dark red dash-dotted curves). The simulations were performed for (a) Brazil, (b) SP, (c) RJ, (d) CE, (e) AM, (f) PA, (g) PE, (h) MA, and (i) BA. The arrows and the percentage indicate the increase or decrease in the number of active infected people *I*(*t*) at the peak of each case when compared with the current trend.

In Figs. 7(a) (Brazil), 7(b) (SP), 7(c) (RJ), 7(e) (AM), 7(f) (PA), and 7(h) (MA), localities for which *ℛ_e_ >* 1.6, we observe an enhancing in the peak of *I*(*t*) of at least 42% when increasing the effective reproduction number in 20%. Besides that, for PE and BA, for which *ℛ_e_*(22) *∼* 1.3, to increase in 20% such quantity can generate a peak with *I*(*t*) at least 120% greater than the current trend. Another interesting case is CE, for which *ℛ_e_*(22) = 0.97. For this federative unit, since the effective reproduction number is smaller than 1, the peak was already reached, and the curve *I*(*t*) is in a descendent way [see the blue continuous curve in Fig. 7(d)]. If *ℛ_e_*(22) increases 20%, from 0.97 to 1.16, a new peak occurs and it can be 672% bigger than the first peak. This result demonstrates the relevance of keeping the effective reproduction number below 1. For all cases, decreasing in 20% the value of *ℛ*_*e*_(22) generates a satisfactory reduction of the peak, at least 47%.

## 4 Summary and conclusions

The present investigation has an apparent limitation, being performed with only data from Brazil (and its federative units), which is one country in the whole world. However, the results obtained from modeling the SARS-CoV-2 epidemic in large countries like Brazil have interesting features that demand to be scrutinized due to the wide territorial, demographic, and infrastructural diversity. Brazil has more than 210 million citizens distributed in 27 federative units which belong to five large regions with completely different cultural and social habits, and economic situations. By scrutinizing the heterogeneous spreading of the COVID-19 outbreak in Brazilian territory, we can understand the epidemic current status in each federative unit and project the evolution of the outbreak in the country. This is important because each federative unit has been dealt with the epidemic in different ways when it comes to containment strategies.

The cumulative number of confirmed cases of COVID-19 until May 30^th^, 2020, is demonstrated for Brazil and its 27 federative units (see Table 1). In Fig. 2, for 10 representative states we observe that, after an initial period with a low incidence of newly infected people, a power-law (*α* + *γt^μ^*) growth of such time series takes place. For each federative unit, we estimated a distinct growth exponent. Brazil is still in the power-law regime, as the federative units studied in Fig. 2, except CE which is achieving a plateau. FD, PA, RJ, and AM have the largest power-law exponents (*μ >* 3.0). Such projections are in complete agreement with the recent results regarding the power-law growth of the cumulative number of infected individuals by the SARS-CoV-2 around the world [11, 12].

Although the power-laws growths with distinct exponents may look similar visually, it is interesting to quantify this similarity. This is performed through the computation of the Distance Correlation (DC) between the data of Brazil and each one of its federative units listed in Table 1, as plotted in colors in Fig. 3. In this figure, we show that the power-law growth for Brazil and for some federative units like SP, RJ, CE, and BA are strongly correlated, which is characterized by *C ∼* 1. For other states as, for example, PR, SE, and TO, in most of the time the value of *C ∼* 0.95, which lead us to conclude that such localities showed a different behavior during the pandemic evolution. In fact, these federative units were not so affected by the pandemic.

By using the SIRD model, it was possible to estimate the epidemiological parameters that best fit the real data in each epidemiological week *k* (EPI week): the infection rate *β*(*k*), the recovery time *T*_rec_(*k*), and the death rate *δ*(*k*). Once these quantities are related by Eq. (13), we estimated the *effective reproduction number ℛ_e_*(*k*) for each EPI week and projected some scenarios based on these values. Our findings suggest that the mean value of *ℛ_e_* for the eight most affected federative units in Brazil is about 2, as shown in Table 3. Besides, our simulations showed that the trend for localities with *ℛ_e_ ∼* 2 is to keep following a power-law growth, while federative units with *ℛ_e_ ∼* 1 are getting the flattening of the curve. Moreover, CE is nowadays the only federative unit for which *ℛ_e_* < 1, and its curve of active infected people *I*(*t*) is in a descendent way [see Fig. 7(d)]. We also observed that, by increasing the value of the effective reproduction number in 20%, the peak of *I*(*t*) not only might be at least 40% greater but it also might occur earlier. Our results show that the only way to flatten the curve is to decrease *ℛ_e_*. In further investigations, we intend to improve our findings considering stochastic perturbations to study how the uncertainty of the official data (due to several reasons) might change such projections. Beyond that, the transmissibility of asymptomatic individuals and the vulnerability conditions of some piece of the total populations also may be considered, mainly in developing countries like Brazil, and in least developed countries. Additionally, the airborne COVID-19 transmission also might be achieved in future studies.

## Data Availability

The data used in this study are openly available in Ref. 30 of the manuscript.

https://covid.saude.gov.br/

## Acknowledgments

C.M. thanks CNPq (Brazil) for financial support (Grant Nos. 304918/2017–2, and 424803/2018–6). The authors also thank MSc. E.L. Brugnago and Dr. M.W. Beims for the fruitful discussions about the model implementation.

## Author contributions

R.M.S., C.F.O.M. and C.M. contributed to the design and implementation of the research, to the analysis of the results and to the writing of the manuscript.

## Competing interests

The authors declare no competing interests.

## Availability of materials and data

The data used in this study are openly available in Ref. [30].

## Notes

### Competing Interest Statement

The authors have declared no competing interest.

### Funding Statement

The authors have declared no external funding was received.

### Author Declarations

The authors have declared no necessary IRB and/or ethics committee approvals.

